# Testosterone and longevity in men and women: A Mendelian randomization study in the UK Biobank

**DOI:** 10.1101/2020.06.18.20134775

**Authors:** CM Schooling, JV Zhao

## Abstract

**Background:** After decades of rising life expectancy, life expectancy in the developed West is currently stagnated and remains shorter in men than women. Very well-established evolutionary biology theory suggests that lifespan trades off against reproductive success, possibly sex-specifically. We examined whether a key driver of reproductive success, testosterone, affected lifespan using a Mendelian randomization study of longevity in the UK Biobank to obtain unbiased estimates, along with control exposures.

**Methods:** We applied published genetic instruments for testosterone to obtain inverse variance weighted estimates of associations with longevity, proxied by survival to (i.e., age at) recruitment, in 167020 men and 194174 women. We similarly obtained estimates for smoking initiation, and absorbate, a marker of vitamin C metabolism, because. We also conducted sensitivity analysis.

**Results:** Overall testosterone was associated with poorer survival (0.10 years younger at recruitment per effect size of testosterone, 95% confidence interval (CI) 0.004 to 0.20). As expected, smoking initiation was also associated with poorer survival (0.37 years younger, 95% CI 0.25 to 0.50), but not absorbate (0.01 years younger, 95% CI −0.09 to 0.11). Sensitivity analysis generally gave a similar interpretation

**Conclusions:** Consistent, with well-established theory, testosterone reduced longevity. Several aspects of a healthy lifestyle (such as a low animal fat diet) and several widely used medications (such as statins, metformin, dexamethasone and possibly aspirin) happen to modulate testosterone. Explicitly designing interventions sex-specifically based on these insights might be a means of addressing stagnating life expectancy and sexual disparities in life expectancy.

---

Sexual dimorphism in lifespan is widely evident, including among humans, contemporaneously and historically.^1,2^ Many reasons have been advanced for shorter life expectancy in men than women, from gendered health and health care seeking behavior^3^ through sex-specific nutrient metabolism and steroid hormones^4^ to sex-specific sexual selection pressures,^5,6^ but few of these have been explicitly exploited as a means of promoting healthy aging in both sexes. As the long-term trend of increasing life expectancy in the West now appears to be stagnating,^7^ reconsidering overlooked targets and their implications for the design of interventions might bear consideration.

An obvious modifiable difference between men and women is levels of sex hormones. Estrogen has been intensively investigated as a protective factor that might explain differences in longevity by sex. Observational evidence concerning benefits of exogenous estrogen in women was very promising.^8^ Large-scale trials of estrogen in women and men were stopped early for harm or lack of benefit.^9^ The discrepant findings have been ascribed to overlooked confounding by socio-economic position in the observational studies.^9^ Promising observational evidence exists for longevity benefits of endogenous testosterone in women^10-12^ and men.^13^ Testosterone in men falls with aging and ill-health,^14-16^ likely generating residual confounding, as health status is difficult to adjust for comprehensively. A trial of the cardiovascular safety of testosterone in women has been successfully conducted (https://clinicaltrials.gov/ct2/show/NCT00612742). A small trial of testosterone in frail older men was stopped early,^17^ making it difficult to interpret. A larger trial of exogenous testosterone on cardiovascular disease is now underway in men (https://clinicaltrials.gov/ct2/show/NCT03518034). Theoretically, the expectation for effects of testosterone on health might be in a similar direction to those found for estrogen because of the well-established Darwinian evolutionary biology trade-off between reproduction, and its drivers, on the one hand, and longevity, on the other hand.^18-20^ However, testosterone in humans could be an exception.

Here to clarify, we conducted a Mendelian randomization (MR) study of the effects of testosterone on longevity in men and women. MR studies take advantage of the random allocation of genetic material at conception to obtain unconfounded estimates.^21^ Longevity studies compare characteristics of survivors to older ages with characteristics of younger people because lifetime harmful factors inevitably become less common with increasing age.^22^ In contrast, observational prospective cohort studies comparing mortality incidence after recruitment are open to selection bias from any deaths prior to recruitment having already depleted the susceptibles23 which may attenuate estimates even to the extent of suggesting no benefit of proven interventions.^24^ As such, longevity studies, take advantage of the changing structure of risk factors with age to obtain estimates of effects on mortality free from selection bias.^25^ Smoking initiation was used as a positive control outcome because smoking is very well-known to be addictive and to substantially reduce longevity.^26^ A marker of vitamin C metabolism, absorbate, was used as a negative control outcome because vitamin C is no longer thought to affect mortality.^27^

## Methods

### Data sources

The UK Biobank recruited half a million people intended to be aged 40 to 69 years from across Great Britain in 2006-10.^28^ Average age is about 57 years and just over half the participants are women. We extracted overall and sex-specific genetic associations with age at recruitment from publicly available UK Biobank summary statistics in white British (http://www.nealelab.is/uk-biobank). These specific summary statistics were generated from linear regression adjusted for sex and the first 20 principal components, after excluding poor quality samples. Where relevant genetic information was not available from these summary statistics, we similarly generated corresponding information from the UK Biobank individual data.

### Exposures

We used published sex-specific genetic predictors of testosterone (125 variants for men and 254 for women).^29^ We used published genetic predictors of smoking initiation (361 variants),^30^ and of vitamin C metabolism (1 variant).^31^

### Outcome

In this longevity study, we used age at recruitment to the UK Biobank as a proxy for survival, as there is no particular reason, apart from prior death (i.e., lack of survival), why genetics should generate differences in willingness to participate by age. So, harmful exposures would be expected to be associated with younger age at recruitment.

### Statistical analysis

We obtained the F-statistics for the genetic instruments using an established approximation (square of genetic variant on exposure divided by its variance).^32^ As F-statistic of 10 or less is usually taken as indicating a weak instrument. We aligned genetic associations on the same allele for exposures and outcome. We included palindromic and non-biallelic genetic variants because the genetic associations with exposures and outcome were obtained in the same study or have been used in the same study. We used proxies, as necessary. We meta-analyzed genetic variant specific Wald estimates (ratio of genetic variant on outcome to genetic variant on exposure) using inverse variance weighting (IVW) with multiplicative random effects. IVW estimates assume balance pleiotropy.^33^ We used estimates with different assumptions as sensitivity analysis. The weighted median has a majority valid assumption and assumes >50% of the genetic variants are valid instruments, while the contamination mixture method has a contrasting plurality valid assumption.^33^ MR-Egger detects pleiotropic effects acting on the outcome other than via the exposure, but assumes that the genetic instruments do not act via confounders of exposure on outcome.^32^ As previously, to avoid pleiotropic effects of sex hormone binding globulin, we used established genetic predictors of bio-available testosterone in men and total testosterone in women.^34^ Bio-available testosterone may also be a more sensitive indicator that total testosterone of active testosterone in men.^35^ Where necessary we combined estimates for men and women using IVW meta-analysis.

We used the MendelianRandomization R package to obtain estimates. We did not adjust for multiple comparisons because this is a hypothesis driven study addressing one question with the use of control exposures, both positive and negative. This study only uses publicly available summary associations from the UK Biobank supplemented by individual level data for information not included in the publicly available summary statistics which was obtained under applicant number 42468. The UK Biobank has ethics approval from the North West Multi-centre Research Ethics Committee. All participants gave informed consent.

## Results

In total 167020 men and 194174 women from the UK Biobank were included. Almost all genetic variants for the exposures were available for the outcome, just two variants for smoking initiation were replaced by highly correlated (r^2^ >0.9) variants (rs2587507 replaced by rs745571, and rs2359180 by rs768576) obtained from LDlink (https://ldlink.nci.nih.gov/). The average F-statistic for testosterone was 128.6 in men and 83.6 in women, and for smoking initiation was 44.9. As expected, smoking initiation was associated with younger age at recruitment (Table 1), corresponding to poorer survival. Similarly, as expected absorbate was unrelated to age at recruitment (Table 1). Overall, testosterone was associated with younger age at recruitment, i.e., poorer survival (Table 1), with slightly larger estimates in men than women (Table S1). Estimates were similar in sensitivity analysis using age at recruitment as a categorical variable (age 60 years or less) (Table S2) and using sensitivity analysis (Table S1).

**Table 1:**
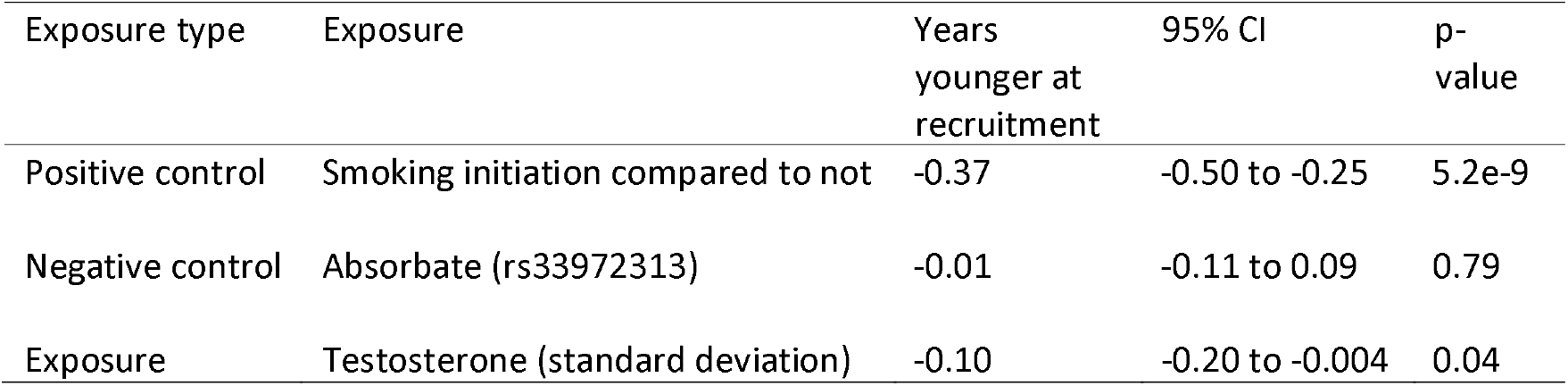
Differences in age at recruitment to the UK Biobank for published genetic predictors of testosterone,^29^ smoking, ^30^ and vitamin C (absorbate)^31^ using Mendelian Randomization inverse variance weighted estimates

| Exposure type | Exposure | Years younger at recruitment | 95% CI | p-value |
| --- | --- | --- | --- | --- |
| Positive control | Smoking initiation compared to not | -0.37 | -0.50 to -0.25 | 5.2e-9 |
| Negative control | Absorbate (rs33972313) | -0.01 | -0.11 to 0.09 | 0.79 |
| Exposure | Testosterone (standard deviation) | -0.10 | -0.20 to -0.004 | 0.04 |

## Discussion

Despite observational studies suggesting that testosterone might promote longevity,^10-13^ our study with greater robustness to confounding and selection bias, consistent with theoretical expectations from evolutionary biology,^18 19^ suggests that testosterone reduces longevity. Our study also showed smoking initiation reduced survival as would be expected.^26^

Our findings differ from previous observational studies which suggest that endogenous testosterone might improve longevity.^10-13^ However, these studies are open to both confounding by health status^14 15^ and selection bias from inevitably only recruiting survivors.^23^ Our findings are more consistent with previous MR studies suggesting exogenous testosterone increases cardiovascular disease,^36,37^ and corresponding warnings from regulators (https://www.fda.gov/drugs/drug-safety-and-availability/fda-drug-safety-communication-fda-cautions-about-using-testosterone-products-low-testosterone-due).

Several potential mechanisms could underlie our findings. Experimental studies suggest testosterone increases coronary plaque volume^38^ and coagulation^39^ while impairing endothelial function.^40-42^ Testosterone increases vulnerability to hormone related cancers, including prostate cancer,^29^ breast cancer^29^ and endometrial cancer.^29^ Testosterone is also increasingly acknowledged to be an immune-suppressant,^43^ potentially increasing susceptibility to cancer,^44^ as well as to infectious diseases,^43^ such as COVID-19. Testosterone also may also induce impulsive behaviour.^45^

### Relevance to interventions

Given the importance of testosterone to reproductive success, testosterone is likely responsive to environmental indicators of suitable conditions for procreation, and hence a modifiable target of intervention. Serendipitously, many aspects of a healthy lifestyle and several widely used interventions reduce testosterone. A healthy diet, i.e., low-fat, high fiber, high soy, and low animal fat reduces testosterone in men^46–48^ and women.^49^ Exercise and weight loss reduce testosterone in women,^50-52^ but possibly less so in men,^53–54^ although severe calorie restriction reduces testosterone in men.^55^ One of the most effective medications for preventing and treating cardiovascular disease, i.e., statins, reduces testosterone in men and women.56 The first-line treatment for diabetes, metformin, may reduce testosterone in women,^57^ but not men.^58^ Aspirin may also reduce testosterone.^59^ Another essential medicine, dexamethasone reduces testosterone.^60–61^ As such, many current interventions to promote longevity do happen to reduce testosterone, particularly in women. Explicitly, searching sex-specifically for interventions based on this insight might facilitate the search for new means of promoting healthy aging, identify at an early stage any potential interventions that are likely to be unsuccessful, and identify where interventions should be sex-specific.

### Limitations

Although this study used an innovative approach to generate an unbiased assessment of the effect of testosterone on longevity, it has several limitations. First, MR has stringent assumptions including that the genetic instruments predict the exposures, hence our use of published instruments,^29-31^ that the instruments are independent of exposure outcome confounders, which stems from the use of genetic instruments not related to confounders such as health status, lifestyle and socioeconomic position, and that the instruments only affect the outcome via the exposure, hence our use of sensitivity analysis and control exposures. Second, we assumed a linear relation of survival with age when it is exponential. However, at the relatively young ages considered here such an approximation is less of an issue than in old age. A sensitivity analysis using survival to age 60 years as the outcome gave a similar interpretation for genetically predicted testosterone (Table S2). Third, the samples used for exposures and outcome overlap. However, that would be expected to bias towards the null,^62^ so our estimates may be conservative. Moreover, the potential bias is less of concern as we did not use weak instruments to predict exposures.^62^ Fourth, our study gives lifetime effects of endogenous testosterone up to age ∼57 years rather than the effect of an intervention. However, the estimates for testosterone can be contextualized by the estimates for smoking. Fifth, longevity studies have been criticized for not taking account of cohort effects.^25^ However, there is no reason to think that genetics are susceptible to cohort effects in the UK Biobank. Sixth, canalization, i.e., compensation for genetic effects might exist. However, this would likely bias towards the null. Seventh, the UK Biobank is not a population representative study. However, we used a study of longevity rather than a traditional prospective cohort study to avoid selection bias. Eighth, the relatively young age at recruitment, and correspondingly high survival means this study lacks power, so we focused on major determinants of longevity, but were not able to ascertain if the effects of testosterone were greater in men than women. Ninth, the effects of testosterone may vary by age. However, causes are generally expected to act consistently, although they may not be relevant in all situations.^63^ Testosterone in men falls with age,^16^ so our findings could be relatively less relevant to older men, but absolute risk of death increases with age.

## Conclusion

Consistent with well-established evolutionary biology theory^18-20^ testosterone appeared to reduce lifespan. Serendipitously, many existing strategies for promoting health reduce testosterone, particularly in women. Explicitly using an established theory as a guide might facilitate the search for new interventions, and draws attention to the importance of sex-specific interventions.

## Data Availability

This study uses data from the UK Biobank that is publicly available or available on application.

## Acknowledgements

We are immensely grateful to all those who contributed to the UK Biobank study. We are also grateful to Ben Neale for making UK Biobank genetic summary statistics publicly available.

## Contributorship

CMS designed the study, conducted the analysis and wrote the first draft. JVZ reviewed and advised on the study design, generated estimates for the individual data from the UK Biobank, checked the analysis and advised on the interpretation.

## Supplementary Tables

**Supplementary Table 1:** Differences in age at recruitment to the UK Biobank for published genetic predictors of testosterone^29^ and smoking^30^ using additional Mendelian Randomization analyses

| Exposure | Method | Subgroup | Years younger at recruitment | 95% CI | P-value | Intercept p | Q (p-value) |
| --- | --- | --- | --- | --- | --- | --- | --- |
| Smoking (361 variants) | WM | all | -0.35 | -0.52 to -0.17 | 0.0001 |  |  |
|  | MR Egger | all | -0.32 | -0.84 to 0.21 | 0.24 | 0.83 | 434.4 (0.004) |
|  | Conmix | all | -0.61 | -0.94 to -0.34 | 0.0003 |  |  |
| Testosterone (125 variants for men, 254 variants for women) | IVW | Men | -0.12 | -0.29 to 0.05 | 0.16 |  |  |
|  |  | Women | -0.09 | -0.22 to 0.03 | 0.14 |  |  |
|  | WM | Men | -0.25 | -0.54 to 0.04 | 0.09 |  |  |
|  |  | Women | -0.28 | -0.51 to -0.05 | 0.02 |  |  |
|  |  | all | -0.27 | -0.45 to -0.09 | 0.003 |  |  |
|  | MR-Egger | Men | -0.20 | -0.48 to 0.07 | 0.15 | 0.47 | 154.2 (0.03) |
|  |  | Women | -0.13 | -0.35 to 0.10 | 0.27 | 0.72 | 262.1 (0.32) |
|  |  | All | -0.16 | -0.33 to 0.02 | 0.08 |  |  |
|  | Conmix | Men | -0.34 | -0.59 to -0.01 | 0.049 |  |  |
|  |  | Women | -0.12 | -0.28 to 0.05 | 0.16 |  |  |
|  |  | All | -0.17 | -0.32 to -0.02 | 0.03 |  |  |

WM: weighted median, IVW: inverse variance weighted, Conmix: contamination mixture model

**Supplementary Table 2:** Recruitment to the UK Biobank at age 60+ years compared to younger than or equal to 60 years for genetically predicted testosterone^29^ using Mendelian Randomization inverse variance weighting estimates

|  | Odds ratio | 95% CI | p-value |
| --- | --- | --- | --- |
| All | 0.98 | 0.95 to 0.99 | 0.049 |
| Men | 0.97 | 0.93 to 1.01 | 0.13 |
| Women | 0.98 | 0.95 to 1.01 | 0.20 |

